# Unsupervised Discovery of Risk Profiles on Negative and Positive COVID-19 Hospitalized Patients

**DOI:** 10.1101/2020.12.30.20248908

**Authors:** Fahimeh Nezhadmoghadam, Jose Tamez-Peña

## Abstract

COVID-19 is a viral disease that affects people in different ways: Most people will develop mild symptoms; others will require hospitalization, and a few others will die. Hence identifying risk factors is vital to assist physicians in the treatment decision. The objective of this paper is to determine whether unsupervised analysis of risk factors of positive and negative COVID-19 subjects may be useful for the discovery of a small set of reliable and clinically relevant risk-profiles. We selected 13367 positive and 19958 negative hospitalized patients from the Mexican Open Registry. Registry patients were described by 13 risk factors, three different outcomes, and COVID-19 test results. Hence, the dataset could be described by 6144 different risk-profiles per age group. To discover the most common risk-profiles, we propose the use of unsupervised learning. The data was split into discovery (70%) and validation (30%) sets. The discovery set was analyzed using the partition around medoids (PAM) method and robust consensus clustering was used to estimate the stable set of risk-profiles. We validated the reliability of the PAM models by predicting the risk-profile of the validation set subjects. The clinical relevance of the risk-profiles was evaluated on the validation set by characterizing the prevalence of the three patient outcomes: pneumonia diagnosis, ICU, or death. The analysis discovered six positives and five negative COVID-19 risk-profiles with strong statistical differences among them. Henceforth PAM clustering with consensus mapping is a viable method for unsupervised risk-profile discovery among subjects with critical respiratory health issues.

## I. INTRODUCTION

**T**HE Coronavirus Disease 2019 (COVID-19) pandemic outbreak has become a public health emergency of international concern due to the rapid spreading of the SARS-CoV-2 virus over the world. The high mortality risk of COVID-19, between 2% and 20% depending on the availability and quality of medical resources and the economic situation [1-2], is one of the main issues of the pandemic. Another issue is that many recovered patients suffer from long-lasting sequels affecting their life and with possible economic implications [3-4]. Hence, there is a need for the discovery of effective treatments aimed to improve or cure COVID-19 cases and control the effects of the disease.

One of the most important tasks in managing COVID-19 is the identification and characterization of the different risk profiles of infected subjects. The correct characterization of the risk profile of a specific subject plays an important role in the prompt selection of effective treatment for that specific patient. Furthermore, it could be an effective medical decision making for resource allocation, and it may provide vital information to identify and protect the most vulnerable populations [5]. Several works have been done in this risk profiling area. COVID-19 studies have discovered the most important disease severity risk factors such as older age, being male, obesity, smoking, and comorbidities including hypertension, diabetes, cardiovascular disease, and respiratory diseases that could significantly affect the prognosis of the COVID-19 infected subjects [5-11]. Furthermore, Gansevoort et al. found that subjects with chronic kidney disease have a very high risk of COVID-19 mortality [12].

As we have stated, it is essential to identify the high-risk factors, but more importantly, is to have tools that predict the disease severity of at-risk populations; henceforth, various supervised approaches have been suggested to identify the risk factors associated with COVID-19 progression. Univariate and multivariate ordinal logistic regression models have been the most common methods used to model risk factors for the severity prediction of disease in patients with COVID-19 [13], while Ji et al. used multivariate Cox regression to explore the risk factors of COVID-19 that have a greater risk of developing into the critical or mortal condition [14]. These efforts have been done in different settings or used limited clinical information [15-17]. Furthermore, supervised approaches are limited because there are many possible risk factors that can be associated with outcome severity, and each risk factor and their combination create a large variety of possible COVID-19 risk-profiles hence a very large data set is required to accurately train complex statistical models.

To address the risk-factor combination issue, we propose the use of robust and unsupervised data clustering, to discern the robust patterns in the subject’s risk presentations that can easily be associated with disease severity and outcomes [18]. By identifying the patient’s risk-profiles via clustering we aim to streamline data analysis for treatment decisions.

There are many different data clustering algorithms [19-22]. Among them are statistical clustering strategies [23-25]. They are robust approaches that develop models that describe data adequately, and each model has its explicit factors that aid in data understanding [26-27]. Furthermore, novel algorithmic advances aid in the discovery of robust data clusters from multidimensional data sets. One such method is consensus clustering [28]. Consensus clustering relies on multiple iterations of the chosen clustering method to discover the most reliable partitions from multidimensional data sets. Besides, one of the robust statistical clustering algorithms is Partitioning Around Medoids (PAM) method that intends to find *K* medoids that minimize the sum of the dissimilarities of the observations to their nearest medoid [29].

This study aims is to determine whether unsupervised discovering of risk risk-profiles of COVID-19 and non-COVID-19 patients seeking medical attention may be useful in identifying the set of hospitalized patients that are at higher risk of either: 1) develop pneumonia, 2) require the use of intensive care unit (UCI) or die from the infection. To achieve this goal, we used the Open Mexican Repository that collects, at the patient level, COVID-19 test results, outcomes (pneumonia diagnosis, ICU, death), and known risk factors like age, gender, pregnancy, smoking, obesity, and common comorbidities like hypertension and diabetes among others.

## II. MATERIALS AND METHODS

### A. Data preparation

Preliminary data used in this study was obtained on May 9, 2020 from the COVID-19 Mexican Open Repository published by the General Directorate of Epidemiology of the Mexico government [30]. On June 8th, we updated our dataset to include 128148 subjects with the following variables: patient ID, age, sex, exposure history, obesity, smoking, pregnancy, the type of patient (Ambulatory/Hospitalized), other underlying comorbidities (diabetes, hypertension, cardiovascular disease, Chronic obstructive pulmonary disease (COPD), asthma, immunosuppression, chronic kidney failure, and other diseases), and the ultimate patient outcome (pneumonia, ICU, intubation, and date of death).

Due to the nature of the Mexican COVID-19 sentinel COVID-19 testing strategy [31], we limited our study to hospitalized only subjects. Among hospitalized patients, there were 13367 and 19958 positive and negative COVID-19 patients. Each patient was described with 35 features, but for this study, we focused on the basic set of 13 risk features and three outcomes. Hence the data set had the potential to provide 6144 different risk-profiles per age group.

The descriptive statistics of the selected features and outcomes for hospitalized patients with positive and negative COVID-19 test results are shown in Table I.

**TABLE I.** THE CHARACTERISTICS OF SUBJECTS USED ON COVID-19 MEXICO HOSPITALIZATION DATA SET The values show the number of subjects has characteristics and mean (SE) for Age. The OR was computed with a confidence interval of 95% for positive *vs* negative COVID-19. *, **, and *** denote a small effect size (between 0.2 and 0.5 for Z and between 1.5 and 2 for OR), a medium effect size (between 0.5 and 0.8 for Z and between 2 and 3 for OR), and a large effect size (larger than 0.8 for Z and more than 3 for OR), respectively.

| Feature | Positive COVID | Negative COVID | Effect Size |
| --- | --- | --- | --- |
| Subjects (male ratio) | 13367 (65.75%) | 19958 (55.92%) | OR=1.18 (1.13- 1.22) |
| Age | 53.75 (0.13) | 44.43 (0.16) | Z=0.3* |
| Pregnancy | 58 (0.43%) | 287 (1.44%) | OR=0.3 (0.23- 0.4) |
| Diabetes | 4099 (30.66%) | 5288 (26.50%) | OR=1.16 (1.11- 1.21) |
| COPD | 549 (4.11%) | 1387 (6.95%) | OR=0.59 (0.53- 0.65) |
| Asthma | 335 (2.51%) | 769 (3.85%) | OR=0.65 (0.57- 0.74) |
| Immunosuppression | 370 (2.77%) | 1273 (6.38%) | OR=0.43 (0.39- 0.49) |
| Hypertension | 4313 (32.27%) | 6126 (30.69%) | OR=1.05 (1.01- 1.1) |
| Cardiovascular | 588 (4.40%) | 1433 (7.18%) | OR=0.61 (0.56- 0.68) |
| Obesity | 3307 (24.74%) | 3497 (17.52%) | OR=1.41 (1.34- 1.49) |
| Chronic kidney | 607 (4.54%) | 1488 (7.45%) | OR=0.61 (0.55- 0.67) |
| Smoking | 1251 (9.36%) | 2151 (10.78%) | OR=0.87 (0.81- 0.93) |
| Other diseases | 584 (4.37%) | 1482 (8.63%) | OR=0.59 (0.53- 0.65) |
| **Outcome** |  |  |  |
| ICU | 1596 (11.94%) | 1457 (7.30%) | OR=1.64 (1.52- 1.76)* |
| Deaths | 5610 (41.97%) | 2510 (12.58%) | **OR=3.34 (3.17-3.51)\*\*\*** |
| Pneumonia | 9490 (71.00%) | 11342 (56.83%) | OR=1.25 (1.21- 1.29) |

### B. Initial Statistical Analysis

The selected features of positive and negative groups were analyzed for differences between positive and negative COVID-19 and each group was further described by the difference in recovered and dead patients. The statistical analysis reported the effect size of all features using Cohen’s d (Z) and odds ratio (OR) for continuous and discrete variables respectively [32]. Finally, we computed the frequency of the top 10 main risk risk-profiles observed in males/females with positive/negative test results and stratified into three age groups: young (20<40), middle (40-60), and old adults (>60). In other words, this report will describe the prevalence of the top 120 risk profiles (Fig. 1).

**Fig. 1.**
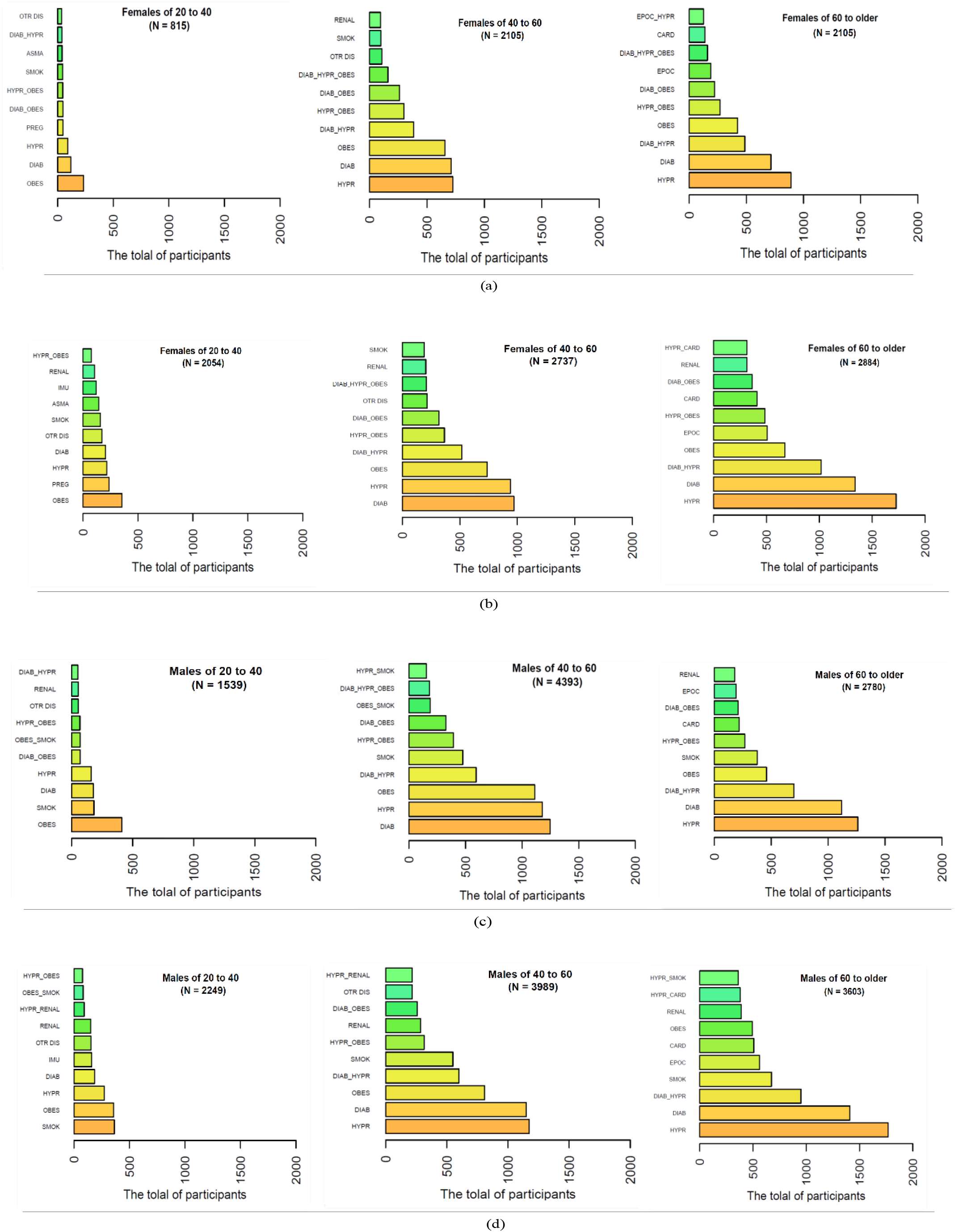
Patients are stratified by sex, age group and COVID-19 test results. The frequencies of the top 10 risk-profiles for, (a) the women of hospitalized infected COVID-19, (b) the women of hospitalized non-infected, (c) the infected hospitalized men, d) the non-infected hospitalized men.

### C. Consensus Clustering and PAM clustering model

Fig. 2 summarizes the overall methodology used for cluster discovering and risk-profile modeling. Firstly, we randomly split the data sets into discovery/training and validation sets. This strategy removes discovery/training biases from the risk evaluation of each patient risk-profile. 70% of the subjects were randomly selected to be part of the cluster discovery and training of the final model for risk-profile prediction. After estimating all the data transformation parameters, the optimal number of clusters, and the final cluster-parameters using the training set, we predicted the corresponding risk-profiles on the remaining 30% of the patients. Finally, the role of each risk characteristic on each one of the risk-profiles was described by Classification and Regression Tree (CART) analysis [33]. All features were standardized between 0 and 1. Age was normalized between the min and max age [34]. Males were coded as one, while females as zero. The rest of the risk categorical features were set to 1 for presence and 0 for the absence of the risk factor. We used the principal components analysis (PCA) transform for dimensionality reduction via the selection of the PCA feature vectors that captured more than 80% of total variance [35-36]. The risk-profile discovery was done as follows. First, we selected the Partitioning Around Medoids (PAM) algorithm as a clustering method [29]. PAM is robust to differences in data distributions, and the user provides the initial *K* medoids. The optimal number of *K*-medoids was found via consensus clustering. Consensus clustering relies on multiple random repetitions of the determined clustering method allowing a robust evaluation of the sensitivity of the clustering approach to input variation [37-39]. Furthermore, the repeated random repetition allows the selection of the *K*-medoids that are more robust to random changes in input parameters. We further enhanced the randomness of the approach by randomly selecting 70% of the subjects for medoid discovery and the holdout discovery samples were used to evaluate the stability of predicting clustering labels on the holdout set. To get a reliable training-holdout-sample clustering evaluation we repeated the procedure 100 times for different values of the number of clusters (*K=2,3,4,5,6,7*). The reliability/stability evaluation of consensus clustering relies on the computation of the cluster co-association matrix (CCAM) [40]. The CCAM is a matrix where each column and row represent a subject in the discovery set, and it stores the counts on how many times two hold-out subjects shared the same cluster label. Hence stable data partitions create a sharp checkerboard pattern, while unstable data partitions create fuzzy patterns. The clarity of the CCAM is analyzed by computing the proportion of ambiguous clustering (PAC). Hence low PAC numbers represent a clarification scheme that is very robust and not sensitive to changes on the discovery set. Henceforth, by repeating the consensus clustering for a variety of different K values the optimal data partition is the one with the lowest PAC number and it represents the most robust and reliable data clustering.

**Fig. 2.**
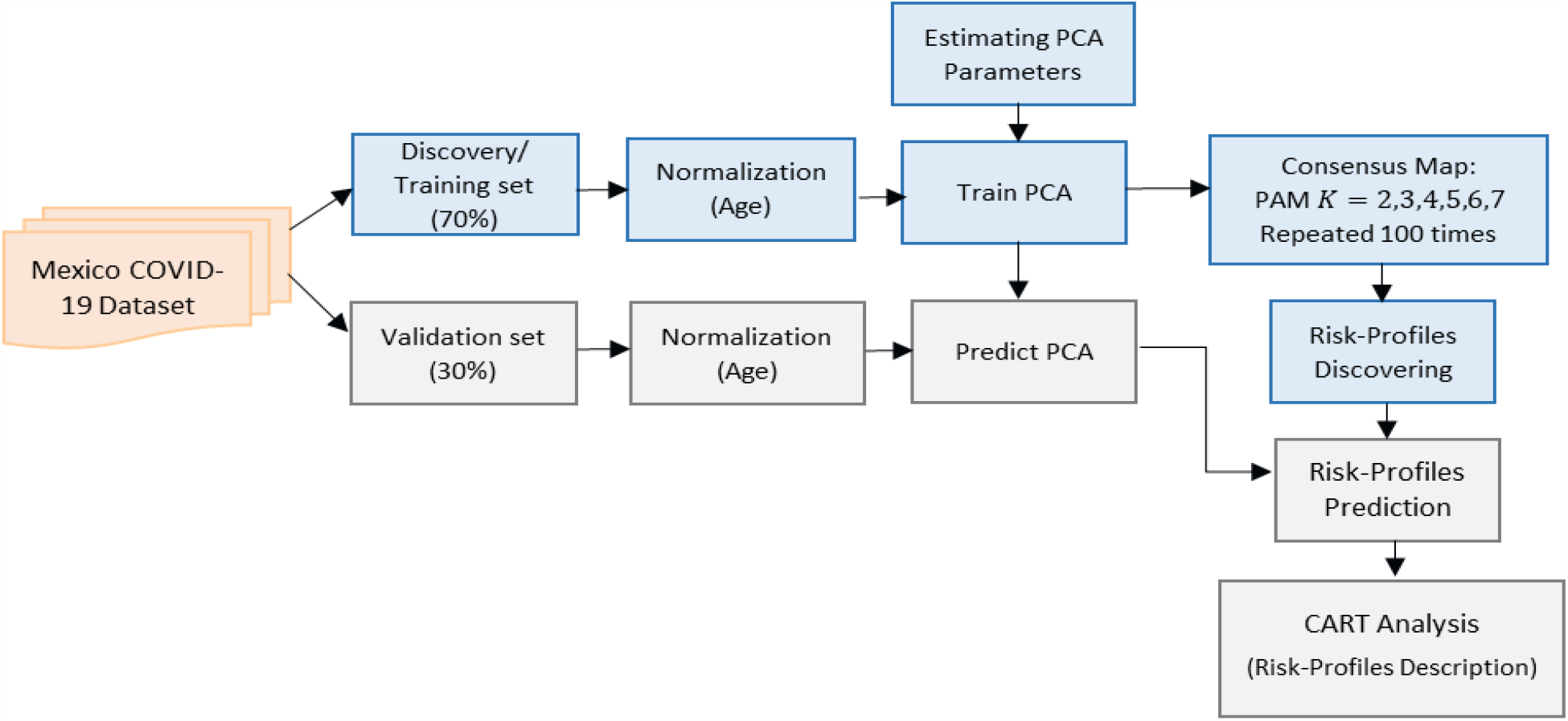
The overall methodology of risk-profile’s classification of Mexico COVID-19 data set. The multimodal data is split into training and testing sets and the results of the testing set are used to describe the association of disease risk-profiles to clinically relevant outcomes.

### D. Statistical and CART Analysis of the discovered risk-profiles

After computing the PCA transform and discovering the best number of clusters and their associated medoid for each discovered risk-profile, we proceeded to predict the risk-profiles of each one of the samples of the validation sets. The risk profile’s prediction is done in three steps: First, normalize the age of the patients. Second, predict the magnitude of each one of the principal components for each subject. Third, label the risk-profiles of the validation sample. This risk-profile prediction returns a unique class label for each subject in the validation set.

After the risk-profile prediction, we analyzed the prevalence of adverse outcomes on each one of the discovered risk risk-profiles. Three adverse outcomes were studied: diagnosis of pneumonia, the requirement of intensive care unit (ICU), or patient death. Consequently, the risk-profile with the highest prevalence of adverse outcomes represents the most critical group. Finally, our final goal was to get simple decision rules for the classification of each new patient into the discovered risk-profiles. For that purpose, we selected the classification and regression trees (CART) analysis. CART automatically creates decision Tree algorithms that can be used for classification or regression predictive modeling problems [41]. Inference regarding the statistical significance of each discovered risk-profile was done either by ANOVA or chi-square test for continuous and discrete values, respectively. Values lower than 0.05 were considered significant, and no effort was made to correct for false discovery.

Implementation and data used are available on GitHub (https://github.com/FahimehN/COVID-19-Risk-Profiles-Discovering).

## III. RESULTS

We first analyzed a cohort of 33,325 hospitalized patients with positive and negative COVID-19 test. Table I illustrates the characteristics of positive and negative COVID-19 hospitalized patients. Statistical differences between them were expressed as effect sizes with 95% confidence intervals (95% CI). The mortality frequency between Positive and Negative COVID-19 was different: OR=3.34 (95% CI= 3.17 to 3.51). In other words, subjects infected with COVID-19 are at a higher risk of death than other patients with other respiratory issues.

Table II shows that age, COPD, chronic kidney, and hospitalization in ICU are moderately different between the people that died and the people that recovered with confirmed COVID-19 test results. Deceased patients were 2.25 and 2.35 times (95% CI, 1.89 to 2.68, and 1.98 to 2.78) more likely to suffer from COPD and Chronic kidney disease compared to the recovered patients, respectively. Moderate effect sizes were observed for age (Z=0.68) and ICU admission (OR of 2.41, 95% CI, 2.16 to 2.68). As such, Table III shows the recovered-death analysis of negative COVID-19 patients. Chronic kidney disease was the strongest risk factor for death. There were marginal differences between the two groups regarding diabetes, COPD, Immunosuppression, hypertension, and Cardiovascular with OR between 1.5 and 2.

**TABLE II.** THE CHARACTERISTICS OF INFECTED SUBJECTS WITH POSITIVE COVID 19 TEST RESULTS BASED ON DEATHS AND RECOVERED. (N= 13367) The values show the number of subjects has characteristics and mean (SE) for Age. The OR was computed with a confidence interval of 95% for dead *vs* alive people. *, **, and *** denote a small effect size (between 0.2 and 0.5 for Z and between 1.5 and 2 for OR), a medium effect size (between 0.5 and 0.8 for Z and between 2 and 3 for OR), and a large effect size (larger than 0.8 for Z and more than 3 for OR), respectively.

| Feature | Deaths | Recovered | Effect Size |
| --- | --- | --- | --- |
| Subjects (male ratio) | 5610 (68.97%) | 7757 (63.43%) | OR=1.09 (1.03- 1.15) |
| Age | 59.33 (0.18) | 49.72 (0.17) | **Z=0.68\*\*** |
| Pregnancy | 9 (0.16%) | 49 (0.63%) | OR=0.25 (0.12- 0.52) |
| Diabetes | 2144 (38.22%) | 1955 (25.20%) | OR=1.52 (1.41-1.63)* |
| COPD | 340 (6.06%) | 209 (2.69%) | **OR=2.25 (1.89- 2.68)\*\*** |
| Asthma | 129 (2.30%) | 206 (2.66%) | OR=0.87 (0.69- 1.08) |
| Immunosuppression | 182 (3.24%) | 188 (2.42%) | OR=1.34 (1.09- 1.65) |
| Hypertension | 2278 (40.61%) | 2035 (26.23%) | OR=1.55 (1.44- 1.66)* |
| Cardiovascular | 347 (6.18%) | 241 (3.11%) | OR=1.99 (1.68- 2.35)* |
| Obesity | 1506 (26.84%) | 1801 (23.22%) | OR=1.16 (1.07- 1.25) |
| Chronic kidney | 382 (6.81%) | 225 (2.90%) | **OR=2.35 (1.98- 2.78)\*\*** |
| Smoking | 565 (10.07%) | 686 (8.84%) | OR=1.14 (1.01- 1.28) |
| Other diseases | 259 (4.62%) | 325 (4.19%) | OR=1.1 (0.93- 1.3) |
| **Outcome** |  |  |  |
| ICU | 1014 (18.07%) | 582 (7.50%) | **OR=2.41 (2.16- 2.68)\*\*** |
| Pneumonia | 4582 (81.67%) | 4908 (63.27%) | OR=1.29 (1.22- 1.36) |

**TABLE III.** THE CHARACTERISTICS OF NON-INFECTED SUBJECTS WITH NEGATIVE COVID 19 TEST RESULTS BASED ON DEATH AND RECOVERED PEOPLE. (N=19958) The values show the number of subjects has characteristics and mean (SE) for Age. The OR was computed with a confidence interval of 95% for dead *vs* alive people. *, **, and *** denote a small effect size (between 0.2 and 0.5 for Z and between 1.5 and 2 for OR), a medium effect size (between 0.5 and 0.8 for Z and between 2 and 3 for OR), and a large effect size (larger than 0.8 for Z and more than 3 for OR), respectively.

| Feature | Deaths | Recovered | Effect Size |
| --- | --- | --- | --- |
| Subjects (male ratio) | 2928 (60.52%) | 17030 (55.11%) | OR=0.54 (0.5- 0.59) |
| Age | 58.05 (0.37) | 46.79 (0.17) | **Z=0.52\*\*** |
| Pregnancy | 3 (0.10%) | 287 (1.68%) | OR=0.06 (0.02- 0.19) |
| Diabetes | 1119 (38.22%) | 4168 (24.47%) | OR=1.91 (1.76- 2.07)* |
| COPD | 309 (10.55%) | 1077 (6.32%) | OR=1.75 (1.53- 2)* |
| Asthma | 84 (2.87%) | 682 (4.00%) | OR=0.71 (0.56- 0.89) |
| Immunosuppression | 279 (9.53%) | 995 (5.84%) | OR=1.7 (1.48- 1.95)* |
| Hypertension | 1222 (41.73%) | 4906 (28.81%) | OR=1.77 (1.63- 1.92)* |
| Cardiovascular | 312 (10.65%) | 1120 (6.68%) | OR=1.69 (1.48- 1.93)* |
| Obesity | 553 (18.89%) | 2941 (17.27%) | OR=1.12 (1.01- 1.23) |
| Chronic kidney | 363 (12.40%) | 1124 (6.60%) | **OR=2 (1.77- 2.27)\*\*** |
| Smoking | 383 (13.80%) | 1770 (10.39%) | OR=1.3 (1.15- 1.46) |
| Other diseases | 328 (11.20%) | 1394 (8.18%) | OR=0.7 (0.62- 0.8) |
| **Outcome** |  |  |  |
| ICU | 362 (12.36%) | 1111 (6.52%) | **OR=2.02 (1.78- 2.29)\*\*** |
| Pneumonia | 2016 (68.85%) | 9248 (54.30%) | OR=1.86 (1.71- 2.02)* |

Moreover, we assessed the prevalence of the top 120 risk-profiles. Fig. 1 displays the frequencies of the top 10 risk-profiles per age/gender and COVID-19 test results. The combinatory analysis results revealed that most men and women 60 years old and over suffered from hypertension or diabetes or both, while obesity is very prevalent in the younger age group between 20 and 40 years old. As such, most hospitalized patients (both males and females with positive and negative COVID) in middle-aged 40 to 60 years old have experienced hypertension, diabetes, or obesity a clear indication that these three comorbidities are clear risk factors for seeking medical care after contracting a respiratory illness.

Afterwards, we discovered the risk-profiles of positive and negative hospitalized COVID-19 subjects on the validation set through consensus clustering and PAM clustering model. Fig. 3 and Fig. 4 illustrate the best CCAM partition and the PAC analysis for the hypothesis of 2 to 7 different risk-profiles for positive and negative COVID-19 subjects, respectively. The best partition for positive COVID-19 patients was composed of 6 clusters, while 5 clusters were present in the negative COVID-19 group.

**Fig. 3.**
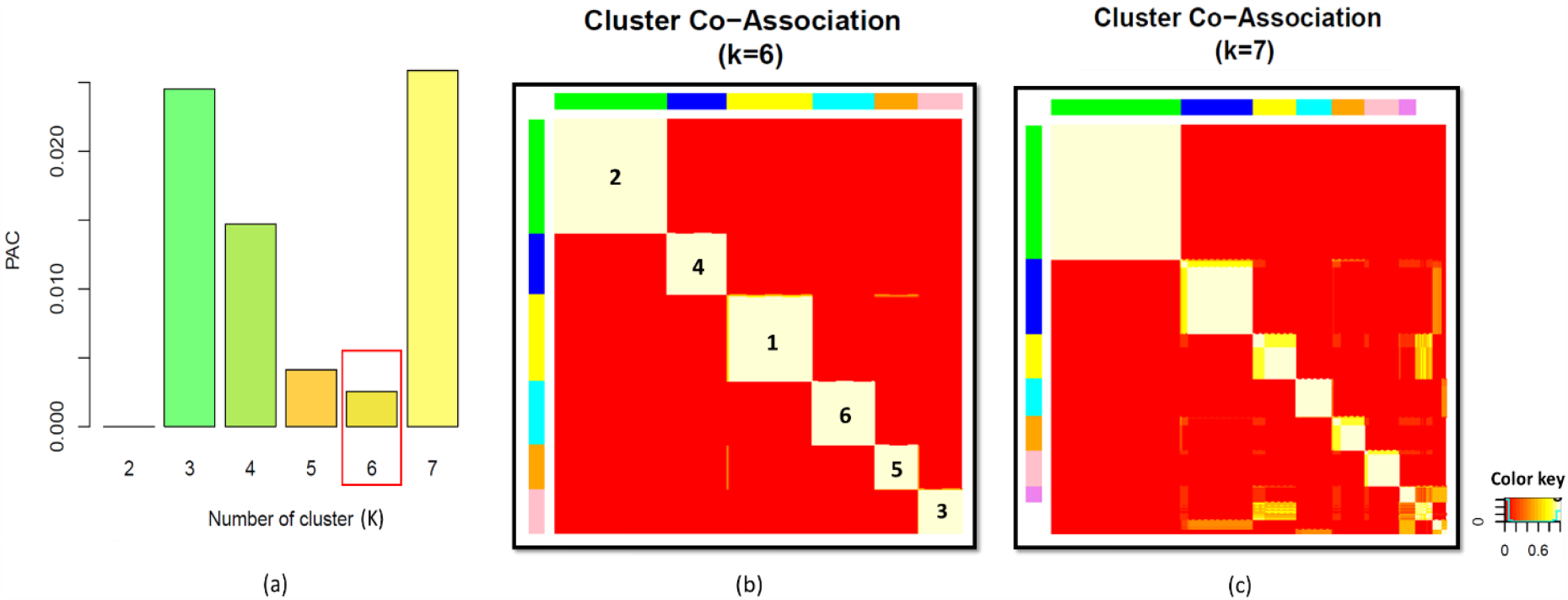
Result of consensus clustering applied to the validation set of subjects with positive COVID-19 test results. (a) The comparison of PAC (smaller is the better) between the cluster numbers from 2 to 7, (b) the best result of Consensus mapping for K = 6, and (c) the worst result of Consensus mapping for K = 7 with the largest value of PAC.

**Fig. 4.**
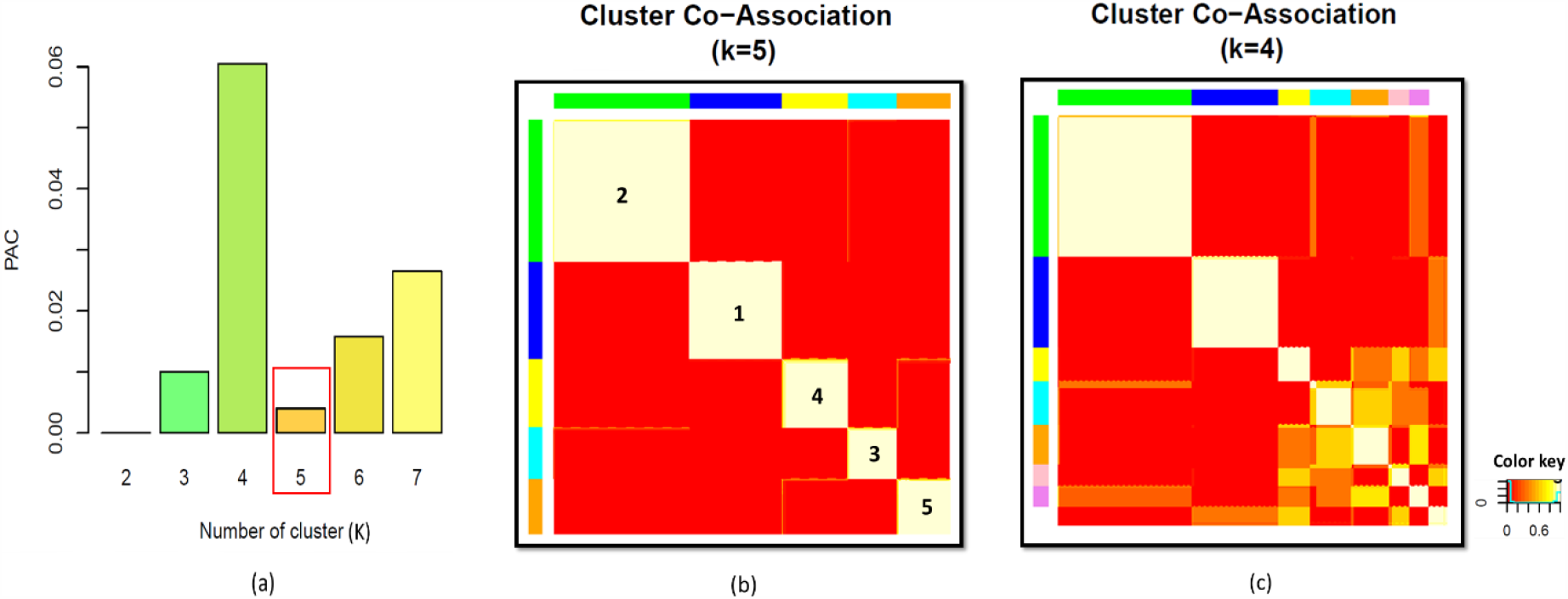
Results of consensus clustering applied to the validation set of subjects with negative COVID-19 test results. (a) The comparison of PAC (smaller is the better) between the cluster numbers from 2 to 7, (b) the best result of Consensus mapping for K = 5, and (c) the worst result of Consensus mapping for K = 4 with the largest value of PAC.

Table IV and Table V show the descriptive statistics of the explored features stratified by risk-profiles for the subjects with positive and negative COVID-19 test results, respectively. Table IV we took the liberty to label three risk-profiles as high death risks (risk-profile 4, 5, and 6). The risk-profile analysis showed that the distribution of features were significantly different values between all risk-profiles. Risk-profile #6 is had the highest risk of death. It was composed mostly of older, males, hypertensive, and diabetic people. Risk-profile #4 had hypertensive subjects without diabetes. While risk-profile #5 was composed of people with diabetes.

**TABLE IV.** THE RESULTS OF THE CLASSIFICATION ON THE VALIDATION SET OF INFECTED PATIENTS WITH POSITIVE COVID-19 TEST RESULTS (6 RISK-PROFILES). The numbers of subjects have the selected characteristics and the Mean (Standard Error) for age were computed in each risk-profile. The p-value was measured by the ANOVA test and chi-squared test between the risk-profiles for continuous and discrete variables, respectively.

| Feature | Low Risk |  |  | High Risk |  |  | p-value |
| --- | --- | --- | --- | --- | --- | --- | --- |
|  | Risk-profile 1<br>(N=678) | Risk-profile 2<br>(N=1174) | Risk-profile 3<br>(N=444) | Risk-profile 4<br>(N=616) | Risk-profile 5<br>(N=429) | Risk-profile 6<br>(N=670) |  |
| Gender (male ratio) | 0 | 1174 (100%) | 348 (78.38%) | 379 (61.52%) | 323 (75.29%) | 386 (57.61%) | P< 0.001 |
| Age | 49.7 (0.56) | 50.15 (0.44) | 44.42 (0.58) | 60.27 (0.58) | 59.06 (0.56) | 61.13 (0.45) | P< 0.05 |
| Pregnancy | 13 (1.92%) | 0 | 0 | 0 | 0 | 0 | P< 0.001 |
| Diabetes | 72 (10.62%) | 0 | 56 (12.61%) | 0 | 429 (100%) | 670 (100%) | P< 0.001 |
| COPD | 29 (4.28%) | 23 (1.96%) | 9 (2.03%) | 45 (7.30%) | 13 (3.03%) | 73 (10.89%) | P< 0.001 |
| Asthma | 22 (3.24%) | 14 (1.19%) | 17 (3.83%) | 18 (2.92%) | 4 (0.93%) | 17 (2.54%) | P< 0.01 |
| Immunosuppression | 31 (4.57%) | 28 (2.38%) | 7 (1.58%) | 14 (2.27%) | 6 (1.40%) | 22 (3.28%) | P< 0.01 |
| Hypertension | 7 (1.03%) | 0 | 6 (0.89%) | 616 (100%) | 0 | 670 (100%) | P< 0.001 |
| Cardiovascular | 9 (1.33%) | 18 (1.53%) | 9 (2.03%) | 45 (7.30%) | 10 (2.33%) | 64 (9.55%) | P< 0.001 |
| Obesity | 98 (14.45%) | 0 | 444 (100%) | 193 (31.33%) | 66 (15.38%) | 211 (31.49%) | P< 0.001 |
| Chronic kidney | 15 (2.21%) | 21 (1.79%) | 94 (1.13%) | 25 (4.06%) | 21 (4.89%) | 69 (10.30%) | P< 0.001 |
| Smoking | 25 (3.69%) | 108 (9.20%) | 58 (13.06%) | 55 (8.93%) | 43 (10.02%) | 68 (10.15%) | P< 0.001 |
| **Outcome** |  |  |  |  |  |  |  |
| Pneumonia | 446 (65.78%) | 809 (68.91%) | 323 (72.75%) | 457 (74.19%) | 332 (77.39%) | 519 (77.46%) | P< 0.001 |
| ICU | 61 (9.01%) | 113 (9.62%) | 59 (13.29%) | 91 (14.77%) | 62 (14.45%) | 90 (13.43%) | P< 0.001 |
| Deaths | 212 (31.27%) | 410 (34.92%) | 169 (38.06%) | 298 (48.38%) | 216 (50.35%) | 367 (54.78%) | P< 0.001 |

**TABLE V.** THE RESULTS OF THE CLASSIFICATION ON THE TESTING SET OF NON-INFECTED PATIENTS WITH NEGATIVE COVID19 TEST RESULTS (5 RISK-PROFILES) The numbers of subjects have the selected characteristics and the Mean (Standard Error) for age were computed in each risk-profile. The p-value was measured by the ANOVA test and chi-squared test between the risk-profiles for continuous and discrete variables, respectively.

| Feature | Risk-profile 1<br>(N=1544) | Risk-profile 2<br>(N=2091) | Risk-profile 3<br>(N=646) | Risk-profile 4<br>(N=940) | Risk-profile 5<br>(N=767) | p-value |
| --- | --- | --- | --- | --- | --- | --- |
| Gender (male ratio) | 0 | 2091 (100%) | 474 (73.37%) | 0 | 767 (100%) | P< 0.001 |
| Age | 38.46 (0.57) | 41.19 (0.51) | 59.12 (0.7) | 64.3 (0.47) | 61.88 (0.47) | P< 0.001 |
| Pregnancy | 83 (5.37%) | 0 | 0 | 0 | 0 | P< 0.001 |
| Diabetes | 20 (1.29%) | 28 (1.34%) | 0 | 745 (79.25%) | 767 (100%) | P< 0.001 |
| COPD | 62 (4.01%) | 81 (3.87%) | 68 (10.53%) | 119 (12.66%) | 79 (10.30%) | P< 0.001 |
| Asthma | 91 (5.89%) | 70 (3.35%) | 18 (2.79%) | 52 (5.53%) | 17 (2.22%) | P< 0.001 |
| Immunosuppression | 97 (6.28%) | 132 (6.31%) | 38 (5.88%) | 60 (6.38%) | 44 (5.74%) | P=0.97 |
| Hypertension | 14 (0.91%) | 0 | 646 (100%) | 690 (73.40%) | 468 (61.02%) | P< 0.001 |
| Cardiovascular | 48 (3.11%) | 73 (3.49%) | 129 (19.97%) | 105 (11.17%) | 73 (9.52%) | P< 0.001 |
| Obesity | 204 (13.21%) | 230 (11.01%) | 171 (26.47%) | 264 (28.08%) | 143 (18.64%) | P< 0.001 |
| Chronic kidney | 22 (1.42%) | 49 (2.34%) | 85 (13.16%) | 125 (13.30%) | 138 (17.99%) | P< 0.001 |
| Smoking | 89 (5.76%) | 255 (12.19%) | 115 (17.80%) | 47 (5%) | 140 (18.25%) | P< 0.001 |
| **Outcome** |  |  |  |  |  |  |
| Pneumonia | 719 (46.57%) | 1168 (55.9%) | 382 (59.13%) | 559 (59.47%) | 505 (65.84%) | P< 0.001 |
| ICU | 117 (7.58%) | 145 (6.93%) | 53 (8.20%) | 62 (6.60%) | 72 (9.39%) | P=0.16 |
| Deaths | 123 (7.97%) | 282 (13.47%) | 121 (18.73%) | 205 (21.81%) | 182 (23.73%) | P< 0.001 |

Table V shows the analysis of the 5 risk profiles of the negative COVID-19 group. Negative COVID-19 subjects had a better chance of survival from their respiratory condition.

It is worth noting that contrary to positive COVID-19 patients, immunosuppression, and requirement for ICU was not significantly different across risk-profiles. The higher risk group was risk-profile #5, where 23.73% of the patients died. It was composed of men (100%) with diabetes (100%) and 61.02% of them experience hypertension.

Fig. 5(a) and Fig. 5(b) represent the results of the CART analysis. The figure depicts the association of risk factors with discovered risk-profiles via decision trees derived from the validation set for positive and negative COVID-19, respectively. Fig. 5(a) shows that 40% of the positive subjects were in high-risk groups (the total of the percentage of observation of risk-profiles 4, 5, and 6 with a higher probability of mortality). The patients who had both hypertension and diabetes were in the highest risk group (Risk-profile 6), whereas the lowest risk group (Risk-profile 1) included the women who did not have hypertension. The analysis of the negative risk profile’s decision trees revealed that there are different decision rules for risk-profiles 4 and 5 and the predicted probability of risk-profiles included mixture values (Fig. 5(b)). Remarkably CART analysis excludes age as significant features for the positive COVID-19 patients.

**Fig. 5.**
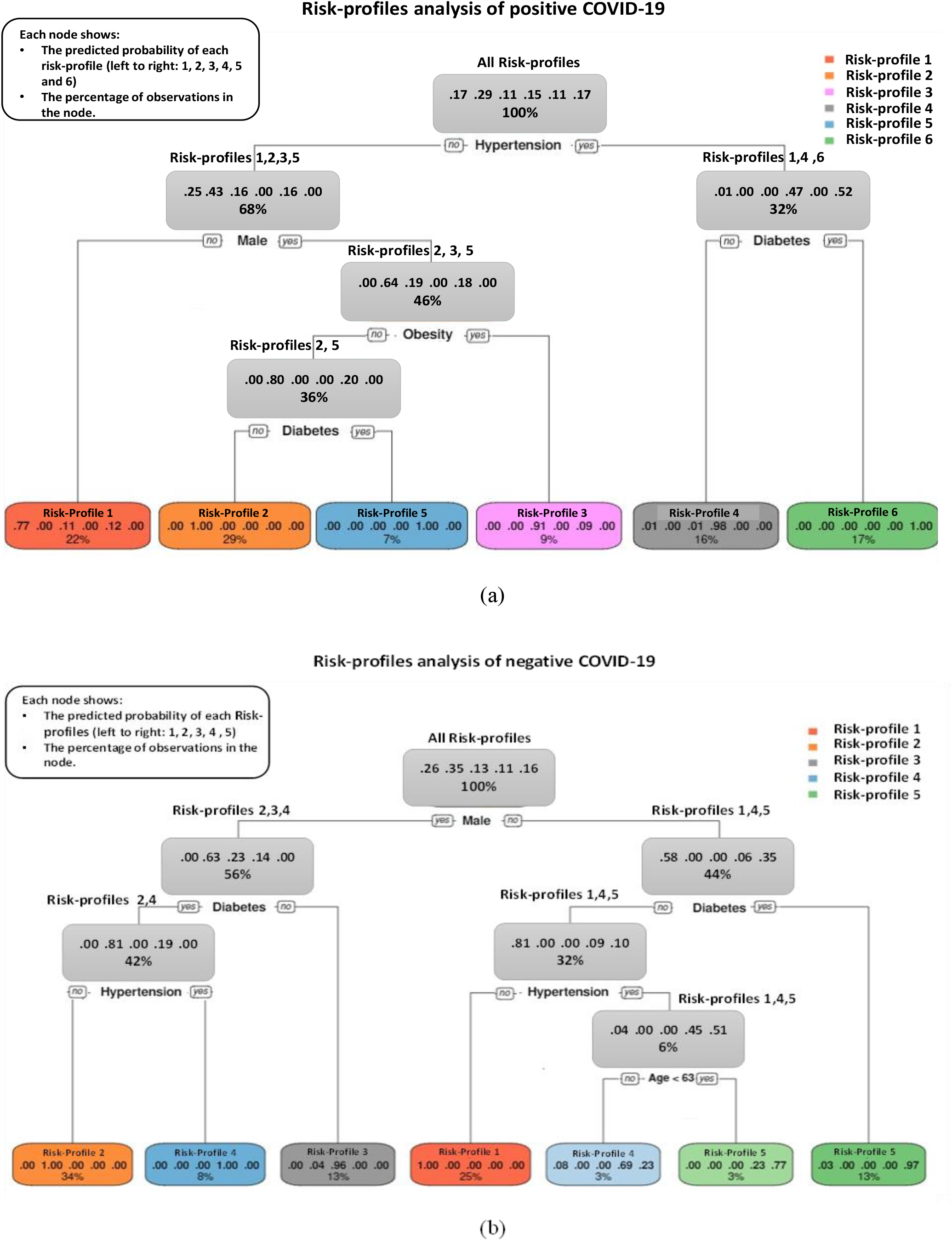
Decision Trees of discovered risk-profiles for, (a) hospitalized infected patients, (b) hospitalized non-infected cases.

## IV. DISCUSSION

In this study, we identified different population risk-profiles of positive and negative COVID-19 hospitalized subjects that were discovered using unsupervised learning via consensus clustering. In the detailed combinatory analysis of the 6144 different risk-profiles per age group that may be presented in the data set, we provided the frequency of the top 10 profiles stratified by gender/COVID-19 test-result/ and age. These results indicated that hypertension, diabetes, and obesity are severely prevalent in 40 years old women. Besides men in this same age group also are more likely to smoke. Smoking and obesity and then diabetes and hypertension are more prevalent for younger men in the 20-40 age group while *smoking* prevalence is less common among women in this age group. These last results are an indication that seeking medical care is strongly associated with health comorbidities confirming previous studies [42-43].

Regarding the main aim of the study of the unsupervised discovery, we found that six and five risk-profiles of infected and non-infected COVID-19 patients were consistently discovered and predicted from the data set. The identification of these risk-profiles was done using a representative training set of the positive and negative COVID-19 groups. The modeling of these risk-profiles with the PAM clustering method enabled us to predict the classes on an independent validation set. Then, the supervised decision trees were used to describe the discovered risk-profiles and extract the decision rules from the discovered risk-profiles.

The severe outcome analysis of the discovered positive COVID-19 groups identified three high risk-profiles. Vulnerable subjects are mostly 60 years or older and with pre-existing medical conditions, such as hypertension, diabetes, and obesity. Also, men are very prone to severe conditions. The analysis of decision rules reveals the highest risk group includes all hypertensive patients with diabetes (risk-profile #6). Other higher-risk groups were mostly men having either hypertension or diabetes (risk-profiles #4 and #5). However, the COVID-19 infected patients in the lowest risk group were women without hypertension (risk-profile #1). We believe that it is important to point out that CART analysis revealed that age was not a discriminant feature for stratifying patients into the six risk-profile groups of COVID-19 patients. Indicating that regales of your age group you are at the same risk and hypertension, obesity, diabetes, and gender are the main factors behind the top six risk-profiles. Our study findings confirm data in previous reports that patients with hypertension and diabetes have more severe illness and higher fatality rates than those without hypertension and diabetes [44-46]. Overall, the identified risk factors for people at increased risk groups are also like those reported in prior studies and showed age, obesity, diabetes, and hypertension are significantly associated with severe COVID-19 [47-49]. In turn, unsupervised clustering models could distinguish patients groups based on a greater risk of severe disease and also can be used to classify newly diagnosed patients that are associated with the risk factors of COVID-19 into known subgroups to facilitate the treatment process.

The analysis of negative COVID subject’s decision rules indicated that some nodes include a mix of risk-profiles without significantly predicted probabilities whereas there were significant differences between more features in different negative risk-profiles. The results showed the highest risk risk-profile of non-confirmed COVID-19 subjects were women with diabetes or hypertensive without diabetes that are older than 63 years old. These imply that the COVID-19 positive patients have more homogenous risk profiles when compared to negative subjects. The hospitalized people who did not get infected with COVID-19 and had negative test results were more likely to prone to other conditions. The disease of these patients who presented symptoms of respiratory can be a bacterial infection, influenza, or other respiratory infections that have a similar disease presentation with COVID-19 [50]. Besides, some of the respiratory symptoms can be related to smoking, however, the percentage of smokers in each negative risk-profile was not significant. On the other hand, the common complication among hospitalized patients with COVID-19 includes severe pneumonia that is a critical lung infection, and may be caused by viral infections, bacterial infections, and other conditions [51]. However, for a few, the coronavirus disease can progress to pneumonia. Moreover, many different sources cause pneumonia and respiratory disorders. Thus, the high risk for pneumonia in negative COVID-19 cases can be associated with other health conditions. Also, the comparison of the outcomes of positive and negative COVID-19 hospitalized cases illustrate that there are significant differences between the mortality and ICU admission rates for both two data sets, and infected COVID-19 patients are more likely to become critically ill and some of them will perish.

An advantage of the clustering method is that the unsupervised clustering models let us predict clusters of patients that were associated with different combinations of risk factors for both positive and negative COVID data sets while supervised decision trees were not able to find the decision rules from the discovered risk-profiles.

This study has several limitations. The findings of this study are based on a Mexican cohort biased toward persons that seeking medical care and hospitalized. Therefore, they can’t be generalized to the overall population. Hence, the findings require validation in an independent cohort. A second limitation is that cluster-based analysis was aimed to discover the main risk profiles, henceforth many different conditions were overlooked, and the simple decision-making rules presented in this study can’t be applied to all subjects. A third limitation is that the outcomes were changing during the pandemic. Different treatments were tested, and hospital saturation varied across patients. Hence, most probably, the risk association results presented in this paper will be only valid to the studied population.

## V. CONCLUSION

This study presented the use the consensus clustering with PAM models to discover the risk-profiles among infected and non-infected COVID patients. We further took advantage of CART analysis to describe the association of discovered risk factors with each risk-profile. Our findings exhibited that the proposed method was able to find a small set of the most common risk-profiles for both data sets, and it may be a useful tool to screen-out the common profiles from other large multi-dimensional datasets. In particular, the results showed that gender, hypertension, diabetes, and obesity are potentially the main high-risk factors for COVID-19 mortality regardless of the age group.

## Data Availability

https://github.com/FahimehN/COVID-19-Risk-Profiles-Discovering

## ACKNOWLEDGMENTS

This research was supported with funding from the Mexican National Council for Science and Technology (CONACYT). The authors are thankful to Dr. Víctor Treviño, Dr. Emmanuel Martínez and Dr. Santiago Conant-Pablos for all valuable comments and suggestions, which helped us to improve the quality of the article.

## Notes

### Competing Interest Statement

The authors have declared no competing interest.

### Clinical Trial

Our study was not a clinical trial

### Author Declarations

Our work did not require ethical oversight.

